# Later Age of Autism Diagnosis in Children with Multiple Co-Occurring Psychiatric Disorders

**DOI:** 10.1101/2025.10.22.25338240

**Authors:** Brian C. Kavanaugh, Danielle G. St. Pierre, Christine Schremp, Asher Robbins, Carrie R. Best, Richard N. Jones, Stephen J. Sheinkopf, Eric M. Morrow

## Abstract

PURPOSE: In children with autism spectrum disorder (ASD), early diagnosis permits early access to therapeutic interventions which may improve outcomes. Factors affecting the age of diagnosis in ASD are not fully understood. METHODS: Here, two large independent datasets were analyzed to investigate age of autism diagnosis and co-occurring psychiatric conditions, including bipolar disorder, depressive disorder, anxiety disorder, obsessive-compulsive disorder, attention deficit hyperactivity disorder, oppositional defiant disorder, and conduct disorder. Clinical characteristics examined included demographics, verbal status, intellectual disability, restricted/repetitive behaviors, adaptive behaviors, and psychiatric medication use. RESULTS: Over 50,000 participants with ASD were analyzed from the Rhode Island Consortium for Autism Research and Treatment study (RI-CART; n=823) and the Simons Foundation Powering Autism Research for Knowledge (SPARK) database (n=52,611). In RI-CART, age of diagnosis differed between those with no co-occurring conditions (mean age at diagnosis = 4.3 years), those with one or two co-occurring conditions (7.1 years), and those with three or more co-occurring conditions (8.5 years; p<.001). This pattern was observed in the SPARK database (age of diagnosis 4, 7.1, and 10 years, respectively; p<.001). Controlling for age, sex, and symptom severity, more co-occurring psychiatric conditions was associated with later age of ASD diagnoses in both samples. Depression and ADHD were associated with later ASD diagnoses; OCD and ID were associated with earlier ASD diagnoses. CONCLUSION: These findings indicate that those children with high co-occurring psychiatric conditions, who are ultimately diagnosed with ASD, experience later diagnosis. This group of children may represent a distinct subtype of autism.

## INTRODUCTION

Autism spectrum disorder (ASD) is a heterogeneous neurodevelopmental disorder characterized by social communication impairments and the presence of restricted, repetitive behaviors (RRBs) (American Psychiatric Association, 2013; Hirota & King, 2023; Lord et al., 2018). ASD often co-occurs with various other conditions (Simonoff et al., 2008; Soke et al., 2018). Although ASD can present as a stand-alone disorder, it is frequently accompanied by intellectual disability (ID), attention deficit hyperactivity disorder (ADHD), epilepsy, or other medical, neurologic or psychiatric conditions that prompt clinical care (Soke et al., 2018). Although estimates vary, the majority of individuals with ASD have at least one co-occurring clinical condition or symptom (Mannion & Leader, 2013; Soke et al., 2018).

For individuals and families seeking clinical care, early diagnosis of ASD allows for timely treatment, which may improve developmental outcomes and adaptive skills when treatment is implemented in early childhood (Zwaigenbaum, 2015). Early intervention can be highly effective and tailored to specific needs, such as focusing on social communication, developing language skills, and reducing behavioral challenges (Zwaigenbaum, 2015). Early diagnosis can benefit caregivers and families by providing access to resources which can alleviate familial stress and parental responsibilities.

These treatments and interventions can also be designed to focus on the specific social and cultural needs of families to provide optimized help for them; however, autism is a highly heterogeneous neurodevelopmental disorder, and efficacy of interventions is variable, potentially due to distinct unrecognized autism subtypes (Geoffray et al., 2025). Autism subtyping is a critical research topic that also has clinical utility. Distinct autism subtypes may lead to different ages of initial ASD diagnosis (Nordahl et al., 2021).

Many factors contribute to differences in the age of ASD diagnosis; notably, males are diagnosed on average of 1.2 years before females (Kavanaugh et al., 2021). This relatively later diagnosis in females may influence developmental outcomes. Potential factors explaining this gap include the presence of a male bias in ASD research, and the generally more subtle presentations of ASD traits in some females (Ochoa-Lubinoff et al., 2023). Early diagnosis has been correlated with prominent speech delays, cognitive or related delays, and/or prominent RRBs. Conversely, the absence of prominent speech or cognitive delays and less pronounced RRBs in females may contribute to delayed diagnosis (Kavanaugh et al., 2021). Previous work in this area shows that individuals with intellectual disability are typically diagnosed earlier in life (Leng et al., 2024; Loubersac et al., 2023; Rattaz et al., 2022), whereas those with ADHD often experience later and potentially delayed diagnosis (Knott et al., 2024; Loubersac et al., 2023; Sainsbury et al., 2023).

The present study set out to explore the relationship between co-occurring conditions and age at diagnosis, among other contributing factors, in two large samples of participants with ASD. Our analysis highlights a significantly later age of diagnosis in children with co-occurring psychiatric conditions. This robust and seemingly generalized finding underscores the need for careful consideration in the diagnosis and care of children with ASD and other co-occurring neuropsychiatric conditions.

## METHODS

### Datasets

In order to further understand the role of psychiatric co-occurring conditions on the age at ASD diagnosis in the clinical setting, data were analyzed from two large and independent studies, the Rhode Island Consortium for Autism Research and Treatment (RI-CART) (McCormick et al., 2020) and the Simons Foundation Powering Autism Research for Knowledge (SPARK) (SPARKConsortium, 2018). The RI-CART study is a community-based, state-wide study with standardized, research level autism diagnosis, and a group of patients with a high number co-occurring psychiatric conditions (McCormick et al., 2020); whereas, the SPARK study represents one of the largest autism studies to-date across the United States (SPARKConsortium, 2018). Both datasets contained similar fields regarding age at diagnosis, co-occurring psychiatric conditions, and other variables of interest, making a parallel analysis appropriate. For ease of comparison, a side-by-side study flowchart is provided here (Fig. 1).

**Figure 1.**
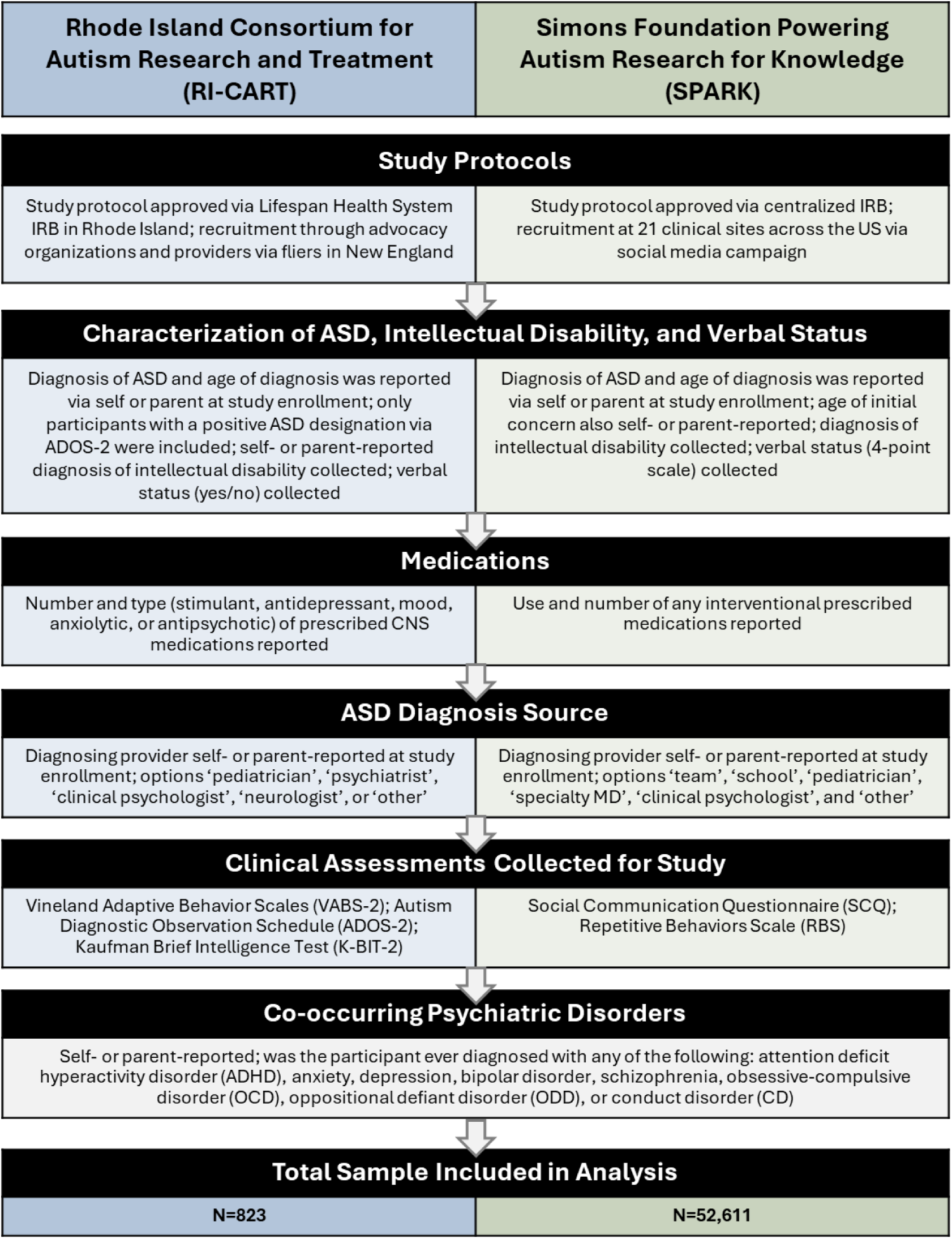
Description of Data Collection Methodology by Study.

### Study Protocols

Both RI-CART (McCormick et al., 2020) and SPARK (SPARKConsortium, 2018) studies were IRB approved and participants and/or guardians provided informed consent. Study protocols for RI-CART were approved via the Lifespan Healthcare IRB (#411409). RI-CART recruited participants through both advocacy organizations and providers. Information for RI-CART recruitment purposes was disseminated through word-of-mouth and social media, along with fliers and presence at community events. Data were collected in Rhode Island and neighboring New England states. Demographics, including sex and race/ethnicity were available.

The SPARK study was approved by a centralized IRB for 21 clinical sites across the United States; consent was received through electronic informed consent forms. Recruitment to SPARK used a multi-pronged social media campaign, with the clinical sites serving as the primary recruitment sites. The dataset included demographic information.

### Characterization of Autism Status and Intellectual Disability

In RI-CART, a previous community-based diagnosis of ASD and the age at that diagnosis was recorded via parental or self-report at study enrollment. In the RI-CART analyses described here, participants were only included if they met the following ASD criteria: a) Enrolled in RI-CART after receiving an ASD diagnosis from a provider in the community and b) Completed the ADOS-2 (Lord et al., 2012) as part of RI-CART study procedures and had an ADOS-2 score reflecting “autism” or “autism spectrum” scoring categories (McCormick et al., 2020). Both prior diagnosis of intellectual disability (‘Yes’/‘No’; ID) and verbal function (‘Yes’ or ‘No’ answers to the query “Is the participant currently verbal?”) were captured via parental or self-report.

In SPARK, prior ASD diagnosis and age at diagnosis were captured via parental or self-report, as well as prior diagnosis of ID. Participants were also asked at what age initial concerns related to ASD were noted. Verbal status was assessed on a 4-point scale from ‘significantly below average’ to ‘above age’ for language level.

### Diagnosing Providers and Medication Use

Both datasets provided information about the source of each individual ASD diagnosis. In both studies, this information was gathered via self or parental-report at the time of enrollment. Information about medication use was also available in both datasets. Differences in the information gathered between datasets are detailed below.

In RI-CART, participants indicated their diagnosis source, given the options ‘pediatrician’, ‘psychiatrist’, ‘clinical psychologist’, ‘neurologist’, or ‘other’. Information on medications which affect the central nervous system was collected, including the type of medication and the number of total medications that have been used. Medication types included stimulant/non-stimulant drugs to treat ADHD and related conditions; antidepressants, such as SSRIs; medications for mood and anti-epileptic drugs (AEDs); anxiolytic medications; and anti-psychotic medication.

In SPARK, options for the source of diagnosis included ‘team’, ‘school’, ‘pediatrician’, ‘specialty MD’, ‘clinical psychologist’, and ‘other’. Medication use was captured by answering ‘yes’ or ‘no’ to the use of any interventional medications.

### Clinical Assessments Collected for Study

Both studies administered clinical assessments related to ASD and provided domain and summary scores as appropriate to the measures. Trained researchers conducted all assessments.

The RI-CART study used the following assessments: 1) the Vineland Adaptive Behavior Scales, Second/Third Edition: Adaptive Behavior Composite (VABS: ABC) (Sparrow, 2005) as a measure of adaptive functioning; 2) the Autism Diagnostic Observation Schedule, Second Edition (ADOS-2) (Lord et al., 2012) as a measure of repetitive, restricted behaviors/stereotyped, restricted interest and social affect/communication, and interaction symptoms; and 3) the Kaufman Brief Intelligence Test, Second Edition (KBIT-2) (Kaufman, 2004) (KBIT-2 administered only to subset of participants). In RI-CART, ASD severity was operationalized using the standardized calibrated severity score, which is calculated using scores from ADOS-2, such that higher scores indicate higher severity (Hus & Lord, 2014; Kavanaugh et al., 2022).

The SPARK study administered: 1) the Social Communication Questionnaire: Total Score (SCQ) (Rutter et al., 2003) as a measure of social communication severity; and 2) the Repetitive Behaviors Scale, Revised Edition (RBS-R) (Bodfish, 2000) as a measure of restricted/repetitive behavior severity. In SPARK, as the ADOS-2 was not available, ASD severity was operationalized by scores on the SCQ consistent with prior work (Memisevic et al., 2023), such that a higher score indicates higher severity. For the SCQ (scores 0-39), a total score of 15 or above is generally considered a clinically meaningful cut-off point suggesting the need for further evaluation (Rutter et al., 2003).

### Co-occurring Psychiatric Disorders

Both studies collected information related to psychiatric diagnoses that co-occurred with ASD. This information was parental or self-reported and captured information about diagnosis with any of the following: attention deficit hyperactivity disorder (ADHD), anxiety, depression, bipolar disorder, schizophrenia, obsessive-compulsive disorder (OCD), oppositional defiant disorder (ODD), and conduct disorder (CD). In RI-CART, the survey item for each condition was “Has the participant received a diagnosis for X?” In SPARK the survey item was “Please select all conditions that (the participant) has been diagnosed with by a professional.” The total number of psychiatric disorders for each person was summed from these categories. While ADHD is sub-categorized as a neurodevelopmental disorder by the DSM-5, for this study ADHD was included among the above described group of psychiatric conditions, whereas ID is not included in the calculation.

### Statistical Analyses

To characterize relationships between the frequency of co-occurring psychiatric conditions and other variables of interest, the cohorts were stratified into the following groups: a) zero co-occurring conditions, b) 1-2 co-occurring conditions, and 3+ co-occurring conditions. These groupings were selected to broadly capture a group with no other conditions, a group with numerous co-occurring conditions, and a middle group with some co-occurring conditions across two separate datasets. Such analyses were done to describe clinical features of these datasets and provide the foundation for the regression analyses. Analyses of variance (ANOVA) and chi-squared analyses examined differences across groups in both datasets. Next, a series of linear regression analyses examined the effect of the total number of psychiatric disorders (i.e., anxiety, depression, bipolar, schizophrenia, OCD, ODD, CD, ADHD, yet excluding ID) diagnosed in the person on the age at ASD diagnosis, after controlling for age at study enrollment, sex, ASD severity, and the presence of ID. Additional regression analyses examined the potential effect of each individual disorder on the age at ASD diagnosis, after controlling for age and sex. Follow-up analyses of covariance (ANCOVA) examined group differences in age at ASD diagnosis between those with and without a given neuropsychiatric disorder, after controlling for age (at enrollment) and sex, allowing estimated marginal means of age at diagnosis to be calculated in the presence or absence of specific disorders. The standardized linear regression coefficient (β), group *F* statistic, and p value are reported. Statistical significance was set at *p* < .05. Data were analyzed in IBM SPSS statistics software.

## RESULTS

### Participant Demographics

Participants enrolled in RI-CART or SPARK and diagnosed with ASD with a known age of diagnosis were included in the analyses. Data from 53,434 individuals were included overall.

In RI-CART (n=823), the mean age at diagnosis was 5.9 years (±6.2). Mean age at study enrollment was 12.6 years (±8.8). Twenty percent of the sample identified as female, 82% selected ‘White’ as their race, and 17% selected Hispanic or Latinx as their ethnicity.

In SPARK (n=52,611), the mean age at diagnosis was 5.8 years (±6.9), while mean age at study enrollment was 11.2 years (±9.1). The mean age of initial concern related to ASD in SPARK was 1.8 years (±1.4). Twenty-five percent of the sample identified as female, 68% selected ‘White’ as their race, and 14% selected Hispanic or Latinx as their ethnicity.

### Clinical Characteristics

In RI-CART, 21% of the sample was diagnosed with intellectual disability (ID), and 85% were reported to be verbal. The most diagnosing providers reported were clinical psychologists (34%), followed by neurologists (20%). The most reported co-occurring psychiatric diagnosis was anxiety disorder (33%), followed by ADHD (30%). Of CNS medications reported, the most frequent were antidepressants (28%), followed by stimulant/non-stimulant ADHD medications (27%).

In SPARK, 21% were diagnosed with ID and 67% were ‘significantly below age’ or ‘below age’ in language level. Here, the most common sources of diagnosis were “team” (50%), and “school” (50%), followed by specialty MD (40%; note: participants could ‘check all that apply’ here). The most reported co-occurring psychiatric diagnosis was ADHD (37%), followed by anxiety disorder (22%). Eleven percent of participants reported use of any interventional medication.

### Group Comparisons between Zero, Moderate, and High Number of Co-Occurring Psychiatric Disorders

Both datasets were stratified into three groups for comparison: no co-occurring conditions (ASD only), moderate number of co-occurring conditions (1-2 psychiatric diagnoses), and high number of co-occurring conditions (3 or more psychiatric diagnoses). Group differences were assessed.

In RI-CART, 51% (n=422) reported no co-occurring conditions, 36% (n=294) had a moderate number of co-occurring conditions, and 13% (n=107) had a high number of co-occurring conditions (Table 1). Mean age of diagnosis was higher in those with 3+ co-occurring conditions (mean age at diagnosis =8.5 years), compared to those with no co-occurring conditions (M= 4.3 years; Cohen’s D = .7), or to those with one or two co-occurring conditions (mean age=7.1 years; *p*<.001; Cohen’s D = .2). Across groups, the ability to speak (verbal status=yes) increased with number of co-occurring diagnoses (76%, 93%, and 98%, respectively; *p*<.001); that is, those with ASD alone were more likely to be non-verbal. The average number of CNS medications also increased in this fashion (mean total # of medications was .53, 1.3, and 1.6, respectively; *p*<.001). The most reported psychiatric diagnoses in the two groups with co-occurring conditions were anxiety disorders (58% of the moderate co-occurring conditions group and 95% of the high co-occurring conditions group were diagnosed with anxiety) and ADHD (54% in moderate co-occurring conditions group and 80% in high co-occurring conditions group were diagnosed with ADHD). Notably, the frequency of diagnosed intellectual disability did not vary significantly between the 3 groups (19%, 23%, and 24%, respectively; p=n.s.). There were no sex differences in number of co-occurring diagnoses in RI-CART.

**Table 1:** Demographic Characteristics and Phenotypic Presentation of RI-CART Participants with Community ASD Diagnosis and Psychiatric Co-Occurring Conditions.

| | N | Total Group | No Co-Occurring Conditions (ASD only) | Moderate # of Co-Occurring Conditions (1-2 Psych Dx) | High # of Co-Occurring Conditions (3+ Psych Dx) | F/ $\chi^2$ | P Value |
| --- | --- | --- | --- | --- | --- | --- | --- |
|  |  | N=823 | n=422 | n=294 | n=107 |  |  |
| Age at Enrollment, M (SD) | 823 | 12.6 (8.8) | 10 (8.1) | 15 (9.2) | 16 (7.7) | 41 | <.001 |
| Age of ASD Diagnosis, M (SD) | 737 | 5.9 (6.2) | 4.3 (5.1) | 7.1 (6.5) | 8.5 (7.4) | 29 | <.001 |
| Female, n (%) | 823 | 164 (20) | 89 (21) | 50 (17) | 25 (23) | 2.7 | .256 |
| Hispanic or Latinx, n (%) | 770 | 129 (17) | 82 (19) | 31 (11) | 16 (15) | 11 | .004 |
| White, n (%) | 782 | 640 (82) | 308 (78) | 244 (87) | 88 (85) | 17 | .082 |
| **Clinical Characteristics** |  |  |  |  |  |  |  |
| Intellectual Disability n,(%) | 823 | 175 (21) | 82 (19.4) | 67 (23) | 26 (24) | 1.8 | .398 |
| Verbal “Yes” n,(%) | 781 | 665 (85) | 292 (76) | 270 (93) | 103 (98) | 54 | <.001 |
| ADOS-2 RRB, M (SD) | 814 | 3.8 (2.1) | 4.3 (2.1) | 3.5 (2.0) | 2.8 (1.7) | 31 | <.001 |
| ADOS-2 SA, M (SD) | 814 | 11.7 (4.1) | 13 (3.9) | 11 (4.4) | 10 (3.7) | 22 | <.001 |
| VABS-II ABC, M (SD) | 689 | 67.3 (17) | 67 (16.5) | 68 (18) | 69 (17) | 0.76 | .469 |
| KBIT-2 IQ, M (SD) | 220 | 84.6 (28) | 74 (29.0) | 91 (26) | 94 (24) | 12 | <.001 |
| Total CNS medications, M (SD) | 823 | .94 (1.2) | 0.53 (1.1) | 1.3 (1.2) | 1.6 (1.3) | 66 | <.001 |
| Stim./non-stimulant, n (%) |  | 225 (27) | 53 (13) | 117 (40) | 55 (51) | 101 | <.001 |
| Antidepressant, n (%) |  | 229 (28) | 54 (13) | 119 (40) | 56 (52) | 103 | <.001 |
| Mood and AED, n (%) |  | 67 (8) | 32 (7.6) | 21 (7.1) | 14 (13) | 4.1 | .131 |
| Anxiolytics, n (%) |  | 39 (5) | 13 (3.1) | 18 (6.1) | 8 (7.5) | 5.6 | .061 |
| Antipsychotic, n (%) |  | 136 (17) | 49 (12) | 60 (20) | 27 (25) | 16 | <.001 |
| Diagnosis Source, n (%) | 656 |  | n=324 | n=242 | n=99 | 25 | .002 |
| Pediatrician |  | 96 (15) | 64 (20) | 25 (10) | 7 (8) |  |  |
| Psychiatrist |  | 118 (18) | 42 (13) | 58 (24) | 18 (21) |  |  |
| Clinical Psychologist |  | 221 (34) | 105 (32) | 87 (35) | 29 (34) |  |  |
| Neurologist |  | 133 (20) | 74 (23) | 41 (17) | 18 (21) |  |  |
| Other |  | 88 (13) | 39 (12) | 36 (15) | 13 (15) |  |  |
| Diagnoses, n (%) | 823 |  |  |  |  |  |  |
| Bipolar disorder |  | 23 (3) | 0 (0) | 6 (2.0) | 17 (16) | 80.3 | <.001 |
| Depressive disorder |  | 95 (11.5) | 0 (0) | 33 (11) | 62 (58) | 281 | <.001 |
| Anxiety disorder |  | 273 (33) | 0 (0) | 171 (58) | 102 (95) | 478 | <.001 |
| OCD |  | 90 (11) | 0 (0) | 39 (13) | 51 (48) | 201 | <.001 |
| ADHD |  | 246 (30) | 0 (0) | 160 (54) | 86 (80) | 394 | <.001 |
| Oppositional defiant disorder |  | 47 (6) | 0 (0) | 14 (4.8) | 33 (31) | 152 | <.001 |
| Schizophrenia |  | 2 (.2) | 0 (0) | 0 (0) | 2 (2) | 13 | <.001 |
| Conduct disorder |  | 22 (3) | 0 (0) | 8 (2.7) | 14 (13) | 57 | <.001 |

**Abbreviations:** ASD, Autism Spectrum Disorder; SD, standard deviation; Verbal “Yes”: participant is able to verbally communicate, based on parent report; ADOS-2, Autism Diagnostic Observation Schedule, Second Edition; RRB, Restricted Repetitive Behaviors; SA, Social Affect; VABS-II/III ABC, Vineland Adaptive Behavior Scales, Second Edition: Adaptive Behavior Composite; KBIT-2 IQ: Kaufman Brief Intelligence Test Second Edition; CNS, Central Nervous System; AED, Antiepileptic Drugs; OCD = Obsessive-compulsive disorder; ADHD = Attention deficit hyperactivity disorder

In SPARK, the pattern of results very closely mirrored those from RI-CART. Specifically, 50% (n=27,403) reported no co-occurring conditions, 37% (n=20,079) had a moderate number of co-occurring conditions, and 12% (n=6,756) had a high number of co-occurring conditions (Table 2). Mean age of diagnosis was higher in those with 3+ co-occurring conditions (mean age at diagnosis =10 years), compared to those with no co-occurring conditions (mean age= 4.4 years; Cohen’s D = .8), those with one or two co-occurring conditions (mean age=7.7years; *p*<.001; Cohen’s D = .3). Across analysis groups, the ability to speak (verbal status=yes) increased with number of co-occurring diagnoses (76%, 93%, and 98%, respectively; *p*<.001). Those who reported taking any interventional prescribed medications also increased in this fashion (percentage of those on any interventional medication in each group was 4.8%, 16%, and 21%, respectively; *p*<.001). The most reported psychiatric diagnoses in the two groups with co-occurring conditions were anxiety disorders (32% in moderate group and 85% in high group) and ADHD (69% in moderate group and 89% in high group). A high percentage of the group were females in the 3+ co-occurring diagnoses compared to ASD only and 1-2 diagnoses (32% vs. 24% and 23%, respectively; *p*<.001) in SPARK only. We therefore calculated the size of this sex-by-group across groups for both datasets (Cramer’s V formula), which indicated a RI-CART effect size V = .06 and SPARK effect size V = .07 (both less than a small effect size). We interpret this as comparable across datasets.

**Table 2.**
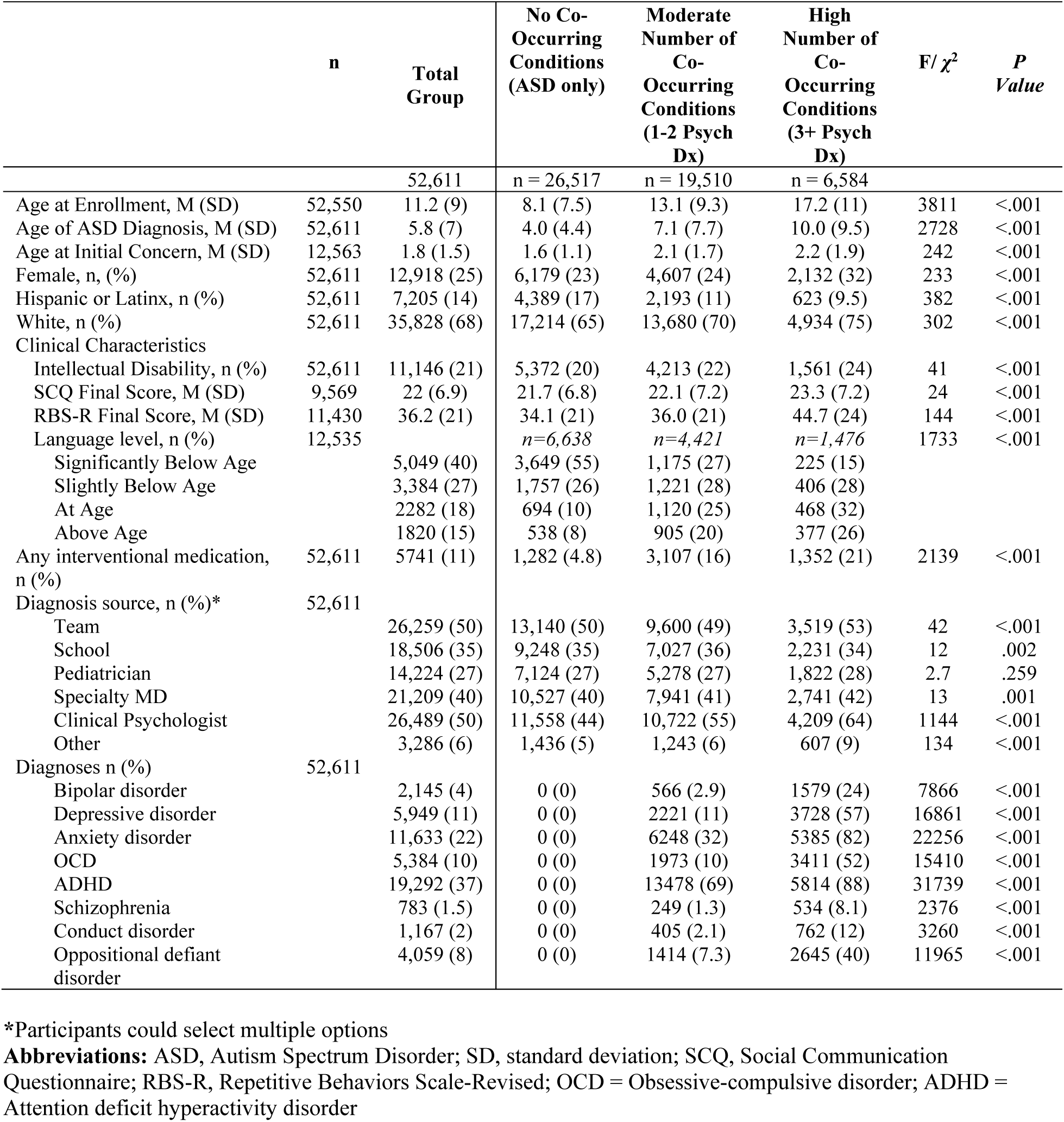
Demographic Characteristics and Phenotypic Presentation of SPARK Participants with Community ASD Diagnosis and Psychiatric Co-Occurring Conditions.

### Age of Autism Diagnosis and Number of Co-Occurring Psychiatric Disorders Regression Models

Group analyses revealed significant differences between zero, moderate, and high number of co-occurring psychiatric conditions with regard to age of ASD diagnosis. Next, we set out to determine the effect of the total number of neuropsychiatric disorders diagnosed in the person on the age at ASD diagnosis using a series of linear regression models controlling for age at study enrollment, sex, ASD severity, and the presence of intellectual disability. Additional regression analyses examined the potential effect of each individual disorder on the age at ASD diagnosis, after controlling for age and sex (Table 3).

**Table 3.**
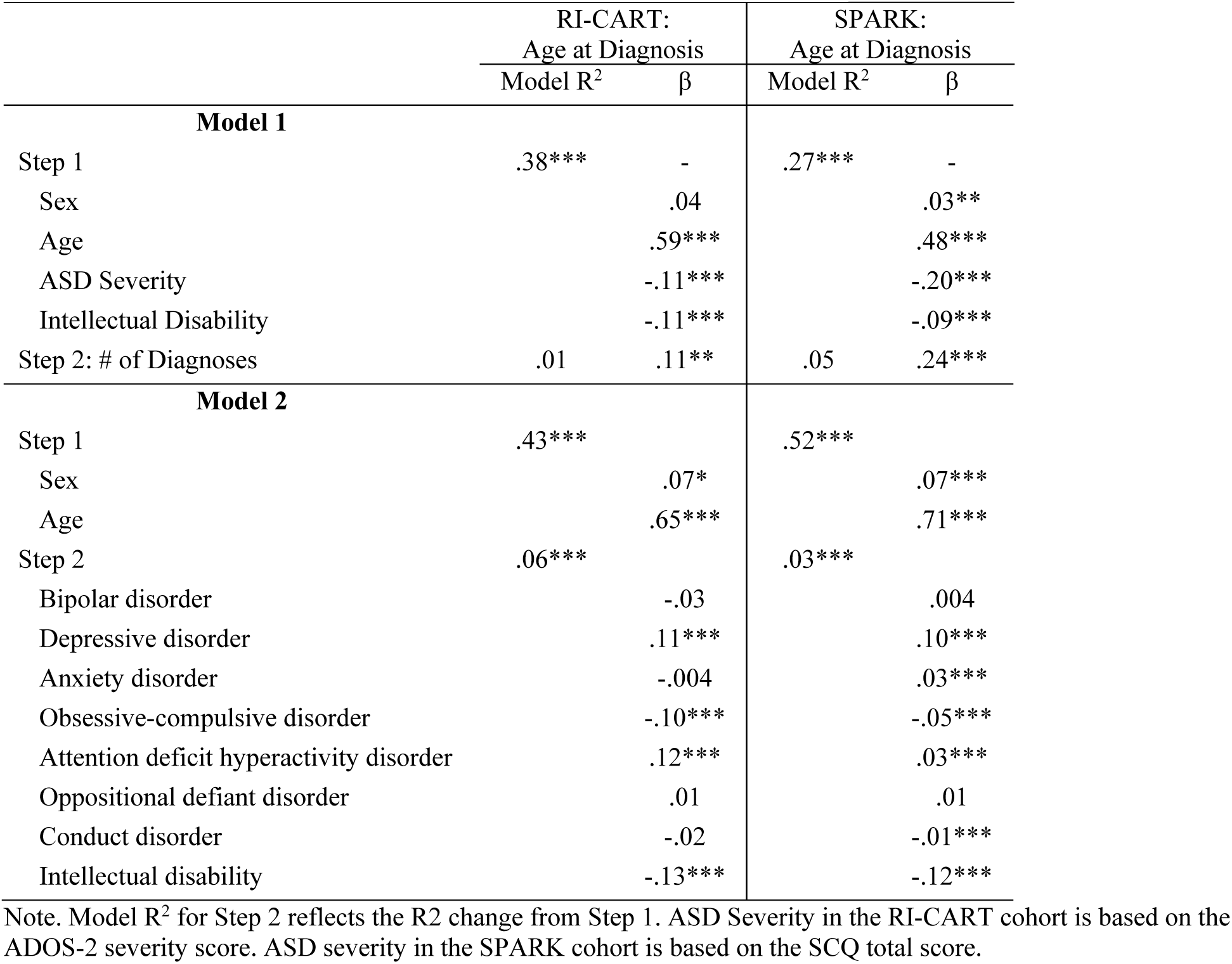
Neuropsychiatric Correlates of Age at ASD Diagnosis.

|  | RI-CART:<br>Age at Diagnosis |  | SPARK:<br>Age at Diagnosis |  |
| --- | --- | --- | --- | --- |
|  | Model R<sup>2</sup> | β | Model R<sup>2</sup> | β |
| **Model 1** |  |  |  |  |
| Step 1 | .38*** | - | .27*** | - |
| Sex |  | .04 |  | .03** |
| Age |  | .59*** |  | .48*** |
| ASD Severity |  | -.11*** |  | -.20*** |
| Intellectual Disability |  | -.11*** |  | -.09*** |
| Step 2: # of Diagnoses | .01 | .11** | .05 | .24*** |
| **Model 2** |  |  |  |  |
| Step 1 | .43*** |  | .52*** |  |
| Sex |  | .07* |  | .07*** |
| Age |  | .65*** |  | .71*** |
| Step 2 | .06*** |  | .03*** |  |
| Bipolar disorder |  | -.03 |  | .004 |
| Depressive disorder |  | .11*** |  | .10*** |
| Anxiety disorder |  | -.004 |  | .03*** |
| Obsessive-compulsive disorder |  | -.10*** |  | -.05*** |
| Attention deficit hyperactivity disorder |  | .12*** |  | .03*** |
| Oppositional defiant disorder |  | .01 |  | .01 |
| Conduct disorder |  | -.02 |  | -.01*** |
| Intellectual disability |  | -.13*** |  | -.12*** |

Note. Model R<sup>2</sup> for Step 2 reflects the R2 change from Step 1. ASD Severity in the RI-CART cohort is based on the ADOS-2 severity score. ASD severity in the SPARK cohort is based on the SCQ total score.

In RI-CART, after controlling for age (β = .59), sex (β =.04), symptom severity (β = -.11), and presence of intellectual disability (ID; β =-.11), the number of co-occurring psychiatric disorders was positively associated with age at ASD diagnosis, in that a higher number of co-occurring psychiatric conditions was associated with a later age at ASD diagnosis (Beta=.11; *p*=.001). We next examined the influence of individual psychiatric disorders on age at ASD diagnosis, after controlling for age (β =.65) and sex (β =.07). Depressive disorders (β =.11, *p*<.001) and ADHD (β =.12, *p*<.001) were associated with an older age at ASD diagnosis. Alternatively, OCD (β =-.10, *p*<.001) and ID (β=-.13, *p*<.001) were associated with a younger age at ASD diagnosis. In RI-CART, no relationship was found between bipolar disorder, anxiety disorder, oppositional defiant disorder, conduct disorder, and age at diagnosis (all *p*=n.s; Table 4.).

**Table 4.**
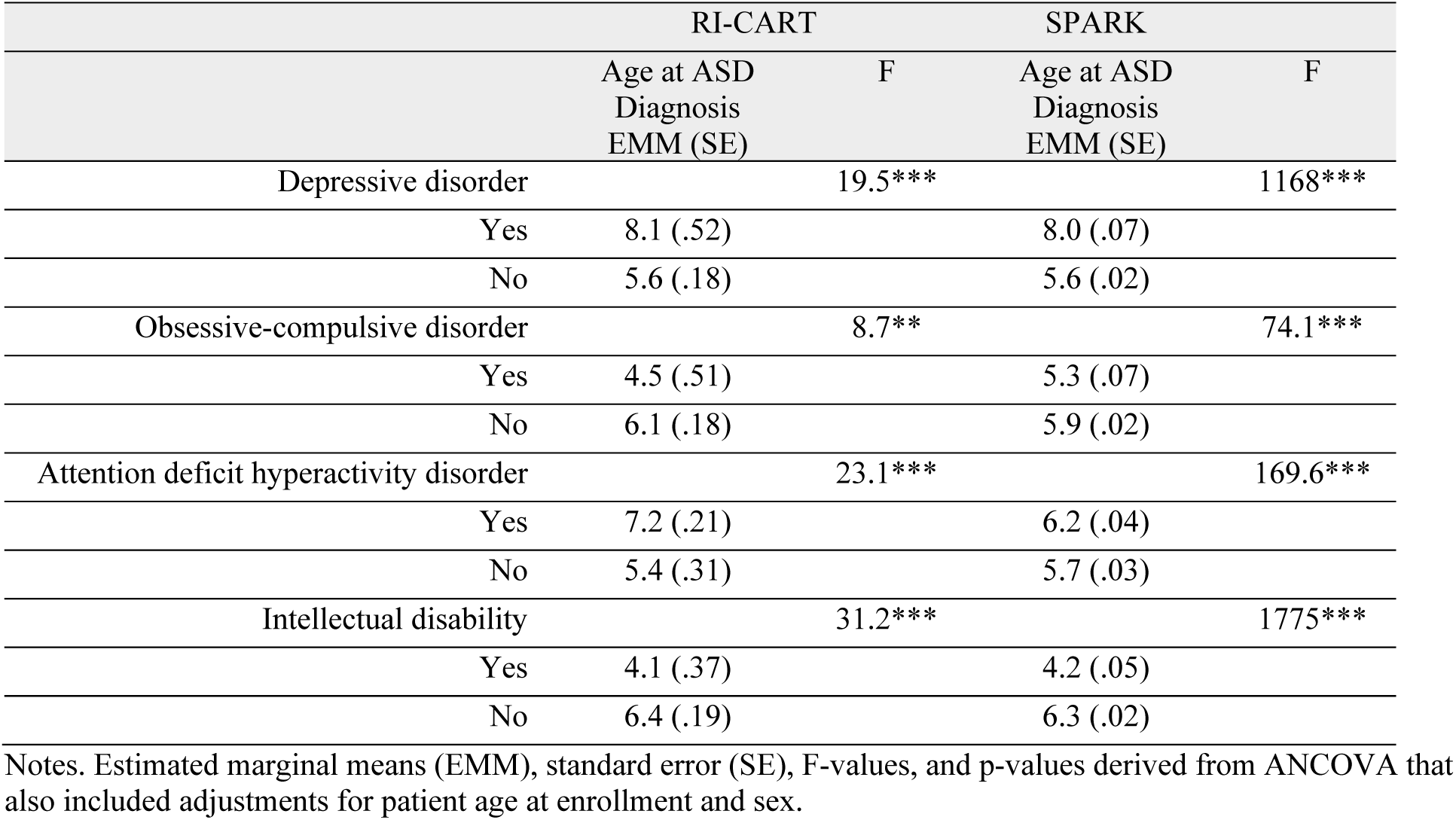
Age at Autism Diagnosis Differences Between Participants with and Without Neuropsychiatric Conditions.

In SPARK, after controlling for age (β =.48), sex (β =.03), symptom severity (β =-.20), and presence of intellectual disability (ID; β =-.09), the number of co-occurring psychiatric disorders was positively associated with age at ASD diagnosis, in that a higher number of co-occurring psychiatric conditions was associated with a later age at ASD diagnosis (β =.24; *p*<.001; Table 3). As in RI-CART, we next examined the influence of individual psychiatric disorders on age at ASD diagnosis within the SPARK dataset, after controlling for age (β =-.71) and sex (β =.07). Depressive disorders (β =.10, *p*<.001), ADHD (β =.03, *p*<.001), and anxiety disorders (β =.03, *p* < .001) were associated with an older age at ASD diagnosis. Alternatively, OCD (β =-.05, *p*<.001) and ID (β =-.12 p<.001) were associated with a younger age at ASD. Conduct disorder was associated with a younger age at ASD diagnosis in the SPARK dataset only (β =.-.01, *p* < .001).

To summarize, linear regression models revealed that depressive disorder and ADHD were associated with a later age at ASD diagnosis across both datasets. Intellectual disability and OCD were associated with an earlier age at ASD in both populations. No association was found between bipolar disorder or oppositional defiance disorder in either dataset. The SPARK dataset, but not RI-CART, showed an association between both anxiety (related to later diagnosis) and conduct disorder (related to earlier diagnosis) and age at ASD diagnosis.

### Age of Autism Diagnosis and Number of Co-Occurring Psychiatric Disorders ANCOVA Tests

To further explore the relationships between specific co-occurring psychiatric diagnoses and age of diagnosis, follow-up ANCOVA analyses examined the age at ASD diagnosis based on presence or /absence of depressive disorders, OCD, ADHD, and ID across both datasets (after controlling for age and sex), as these were the only disorders with significant findings in the linear regression models (Table 4).

In RI-CART, depressive disorders and ADHD were associated with an older age at ASD diagnosis. Specifically, ASD was diagnosed 2.5 years later for those with depressive disorders compared to those without (average diagnosis at 5.6 years old compared to 8.1 years; *F* = 19.5, *p*< .001), and .5 years later for those with ADHD compared to those without (*F* = 23.1, *p*< .001). Conversely, ASD was diagnosed 1.6 years earlier in those with co-occurring OCD (4.5 years old compared to 6.1 years*; F*=8.7, *p*<.001), and 2.3 years sooner in those with intellectual disability (4.1 years old compared to 6.4 years; *F*=31.2, *p*<.001; Table 4).

Again, these results were closely mirrored in the SPARK dataset, such that depressive disorder and ADHD were linked to older age at ASD diagnosis (all *p*<.001) and OCD and intellectual disability were linked to younger age at ASD diagnosis (all *p*<.001; Table 4).

## DISCUSSION

This study, using two large and independent data sets including over fifty-thousand individuals, revealed several important findings related to the age of autism spectrum disorder (ASD) diagnosis and the presence of co-occurring psychiatric conditions. Consistently across both datasets (RI-CART and SPARK), a higher number of co-occurring conditions was associated with a later age of ASD diagnosis. When individual diagnoses were examined, we found that the presence of depressive disorders and attention-deficit/hyperactivity disorder (ADHD) was significantly associated with a later age of ASD diagnosis. However, intellectual disability (ID) and obsessive-compulsive disorder (OCD) were associated with an earlier age of ASD diagnosis. Bipolar disorder and oppositional defiant disorder (ODD) were not significantly associated with variations in the age of ASD diagnosis in either dataset. In the SPARK dataset, anxiety disorders were linked to a later age of diagnosis, whereas conduct disorder was associated with an earlier age of diagnosis; these findings were not observed in the RI-CART dataset.

Our findings indicate that a higher number of co-occurring conditions is associated with a later age of ASD diagnosis. This pattern suggests that the presence of multiple diagnoses may complicate the diagnostic process. Individual disorders which most strongly predicted later diagnosis were ADHD and depressive disorder. We found that those with ADHD were diagnosed at ages 6.2-7.2 years old compared to 5.4-5.6 years old in those without ADHD (Table 4). Prior literature in modest sample size studies (commonly n = 100 to 1,000 participants) have also found that the presence of ADHD is associated with a later age of diagnosis (Knott et al., 2024; Sainsbury et al., 2023). The presence of ADHD was associated with more provider visits prior to receiving an ASD diagnosis (and thus, a delayed diagnosis) in the existing literature (Sainsbury et al., 2023). Notably, ADHD is the only co-occurring psychiatric condition included in this analysis which is also classified as neurodevelopmental. We also found that those with depressive disorder were diagnosed at age 8 years old on average, compared to age 5.6 years in those without (Table 4). Therefore, each disorder alone carries a risk of being diagnosed approximately two years later than non-affected counterparts.

There are various possible mechanisms which may contribute to the associations that we are reporting in this study. ADHD and depressive disorder could possibly mask ASD traits or otherwise lead to diagnostic delays. For example, depressive symptoms, such as withdrawal, may overshadow core social and communication deficits in ASD. Similarly, ADHD-related behaviors such as hyperactivity, impulsivity, and inattention might draw a clinicians attention as the primary and most immediate concern, possibly diverting attention from the more subtle aspects of ASD. Alternatively, depression and/or ADHD may be more common in milder and later emerging ASD. It is also possible that ASD is over diagnosed within the context of comorbid psychopathology, as suggested by others (Fombonne, 2023), or that neuropsychiatric comorbidity can lead to an ASD presentation (Hawks & Constantino, 2020).

In only one of the datasets (SPARK), anxiety disorders were also linked to a later ASD diagnosis. Anxiety may present subtly and can exacerbate social challenges, potentially leading to delayed recognition of core ASD traits or alternatively, leading to a false positive ASD diagnosis (Capriola-Hall et al., 2021). Interestingly, the three disorders linked to delayed diagnosis also were the three most common co-occurring conditions across all participants (37% ADHD, 22% anxiety disorder, and 11% depressive disorders). Anxiety, in particular, was very frequently reported in the groups with 3+ co-occurring conditions (95% in RI-CART and 82% in SPARK), indicating that anxiety is often diagnosed with at least two other conditions. This high incidence could have contributed to the predictive relationship between anxiety and later ASD diagnosis.

Conversely, earlier ASD diagnosis was linked with intellectual disability and OCD. Those with intellectual disability were estimated to be diagnosed around 4.1 years old, compared to those without, diagnosed at 6.4 years old. And, those with OCD were estimated to be diagnosed with ASD between 4.5-5.3 years old, compared to around 6 years old in those without OCD. Different mechanisms may underlie these relationships. Intellectual disability has long been recognized as a strong predictor of earlier diagnosis, likely due to its profound impact on developmental milestones, prompting earlier evaluations. Similarly, the heightened severity and specificity of OCD symptoms, which includes behaviorally manifested repetitive behaviors (Jiujias et al., 2017), in young children with ASD may lead to earlier parental and/or clinical concern related to neurodevelopment. In only one of the datasets (SPARK), conduct disorder was also associated with earlier diagnosis. This could reflect the disruptive nature of conduct disorder on daily life, including symptoms such as aggression and destructive behavior, which may prompt earlier evaluations.

Interestingly, no significant associations were found for bipolar disorder or oppositional defiant disorder in either sample. This may reflect the relatively low prevalence of these conditions in ASD populations: combined across datasets, only 4.1% of participants were diagnosed with bipolar disorder and 7.7% with ODD. Another mechanism could be that these conditions generally present later in development, reducing their influence on the timing of ASD diagnosis. More broadly, we noted numerous consistencies across the two independent datasets, highlighting the reproducibility of current findings. At the same time, there were some differences between RI-CART and SPARK in the strength of relationship between variables of interest. While there are many explanations for these differences (some of which are described below as it relates to measures), RI-CART was a community-based and state-wide study in Rhode Island and SPARK was a nationwide registry involving 20+ clinical sites in the US. Future work should examine possible causes of discrepancies across large-scale studies to maximize data consistency.

Taken together, these findings have significant clinical implications. Although literature on older ages and ASD that emerges later in development remains unclear, an early childhood diagnosis is associated with improved outcomes (Zwaigenbaum, 2015). Our findings raise questions about whether earlier ASD diagnosis is achievable in children with psychiatric co-occurring conditions, and what strategies could be implemented to facilitate this. One possibility is that a care provider may defer a diagnosis of ASD for those with co-occurring conditions to focus on what they perceive as more urgent co-occurring conditions, rather than diagnosing and treating ASD concurrently. Another possibility may be that when co-occurring with other psychiatric conditions, ASD may present atypically or with less overt symptoms, causing diagnostic difficulty. For example, individuals with greater number of co-occurring psychiatry conditions appear to have higher cognitive and adaptive function, fewer language delays, and less outwardly visible or notable repetitive behaviors. Related, individuals with ID or limited verbal skills may still experience these internal mood states but might not be able to as fluently express their internal state and subsequently be diagnosed with a psychiatric condition (Dell’Armo & Tassé, 2024). This extends to the ‘profound autism’ presentation, as the ASD diagnosis and possible ID may be labelled, but other difficult to identify neuropsychiatric co-occurring conditions may be missed in the diagnostic process in this subgroup. Finally, ASD may not fully manifest until later in life as co-occurring conditions evolve or as social demands increase during school-age years. It is possible that the ASD symptoms are subthreshold at earlier ages in individuals with other conditions. Discerning between these different hypotheses could have important clinical implications, particularly when an earlier diagnosis may help navigate treatments and improve outcomes.

Another area wherein this research may have utility is in the recognition of potentially clinically useful subtypes. Autism is a highly heterogeneous neurodevelopmental disorder and recognition of clinically meaningful subtypes is critical to precision care (Nordahl et al., 2021). In this study, we identify a group of people with autism who have a high rate of co-occurring psychiatric conditions and a later diagnosis. This subtype is clearly distinct from the group without co-occurring conditions and early diagnosis. For example, the subgroup with late diagnosis had a higher IQ, were more likely to be verbal and to receive medications.

These findings should be interpreted in the light of limitations. This study examines statistical associations and does not establish causation. Also, importantly, we only have the age of ASD diagnosis, and we do not have the age of diagnosis for the other conditions, so we cannot examine the relationship between age of ASD diagnosis and age of diagnosis of other conditions. Additionally, these data are cross-sectional, which limits the ability to fully describe the timing of other diagnoses and the relationship to ASD symptoms and diagnosis. Longitudinal studies tracking participants over time would help to reveal some of these differences. Another limitation is that, while many variables are comparable or even identical between the two datasets, some did not match well (for example, verbal status as yes/no vs. language ability on a 4-pt scale). Additionally, different clinical assessments were used between the two studies. This could limit the generalizability of the findings. Most variables, particularly symptom severity forms and recall of prior diagnostic history, were largely based on parental reports during a research study visit. Neither RI-CART nor SPARK included within study procedures a diagnostic process to establish a confirmatory ASD diagnosis, so confirmation of ASD diagnostic criteria in participants cannot be made. This may also explain some of the inconsistencies within and across datasets, particularly as it related to co-occurring diagnoses. Further, it is likely that co-occurring psychiatric diagnoses are underdiagnosed in individuals with ID or limited verbal skills given their reduced ability to freely report on their internal mood state. Despite these potential limitations, the strong effect sizes, and remarkable consistency of findings across both large datasets, reflect strengths of this study.

In conclusion, this study contributes to the growing body of literature on ASD diagnosis and co-occurring psychiatric conditions, now in a large dataset. By understanding autism subtypes associated with age at diagnosis, we may be able to inform targeted screening and intervention efforts to prevent delayed diagnoses, tailor treatments to specific subtypes, and potentially improve outcomes for individuals with ASD and their families.

## Data Availability

Approved researchers can obtain the SPARK population dataset described in this study (https://www.sfari.org/resource/spark/) by applying at https://base.sfari.org.

## AUTHOR STATEMENTS

### Conflict of Interest Disclosures (includes financial disclosures)

The authors report no biomedical financial interests or potential conflicts of interest.

### Funding/Support

This work was supported by the Simons Foundation Autism Research Initiative (286756 and 454555), the Norman Prince Neurosciences Institute at Lifespan, and the Robert J. and Nancy D. Carney Institute for Brain Science at Brown University. This work was also supported by the following grants: NIH grants (R01NS113141, R01NS121618, R01MH137004, and R01AG087455) to EMM. BK is supported by K23MH129853.

### Author Contributions

BCK provided substantial methodological and analytic contributions and led the drafting and reviewing/editing of the manuscript; DGS, CS, and CRB provided substantial analytic contributions and participated in manuscript development. CRB also supervised the investigative aspects of the project. RNJ provided substantial statistical contributions. SJS provided substantial methodological, conceptual, and investigative contributions.AR, RNJ, SJS participated in the reviewing/editing of the manuscript. EMM provided substantial methodological, conceptual, and investigative contributions and participated in the drafting and reviewing/editing of the manuscript.

### Role of the Funder/Sponsor

The funding sources had no role in the design and conduct of the study; collection, management, analysis, and interpretation of the data; preparation, review, or approval of the manuscript; and decision to submit the manuscript for publication.

## Acknowledgments

We are grateful to the families who participated in the RI-CART project; without their generous participation, this work would not be possible. Additionally, we would like to acknowledge Hannah Marsland, BA, Rebecca Bradley, BS, Elaine Clarke, BS, and Molly Goldman, MS, of Emma Pendleton Bradley Hospital, for technical assistance, and Hasmik Tokadjian, MS, of Women & Infants Hospital of Rhode Island, for technsical assistance.

*SFARI Data Use:* We are grateful to the families who participate in SPARK, as well as the SPARK clinical sites and SPARK staff. We appreciate obtaining access to the phenotypic data on SFARI Base. Approved researchers can obtain the SPARK population dataset described in this study (https://www.sfari.org/resource/spark/) by applying at https://base.sfari.org.

## Notes

### Competing Interest Statement

The authors have declared no competing interest.

### Author Declarations

Ethics committee/IRB of Lifespan Healthcare (Providence, RI) gave ethical approval for study #411409.

## REFERENCES

American Psychiatric Association. (2013). Diagnostic and statistical manual of mental disorders (5th ed.). American Psychiatric Publishing.

Bodfish, J. W., Symons, F. J., Parker, D. E., & Lewis, M. H. (2000). Repetitive Behavior Scale– Revised (RBS-R) [Database record]. APA PsychTests. 10.1037/t17338-000

Capriola-Hall, N. N., McFayden, T., Ollendick, T. H., & White, S. W. (2021). Caution When Screening for Autism among Socially Anxious Youth. J Autism Dev Disord, 51(5), 1540–1549. 10.1007/s10803-020-04642-w

Dell’Armo, K., & Tassé, M. J. (2024). Diagnostic Overshadowing of Psychological Disorders in People With Intellectual Disability: A Systematic Review. Am J Intellect Dev Disabil, 129(2), 116–134. 10.1352/1944-7558-129.2.116

Fombonne, E. (2023). Editorial: Is autism overdiagnosed? J Child Psychol Psychiatry, 64(5), 711–714. 10.1111/jcpp.13806

Geoffray, M. M., Oreve, M. J., Jurek, L., Sonie, S., Schroder, C., Delvenne, V., Manificat, S., Touzet, S., Agathe, J., Mengarelli, F., Natacha, G., Petit, N., Speranza, M., Bahrami, S., Bouveret, L., Dochez, S. L., Auphan, P., Zelmar, A., Falissard, B., . . . Febvey-Combes, O. (2025). Early Start Denver Model effectiveness in young autistic children: a large multicentric randomised controlled trial in two European countries. BMJ Ment Health, 28(1). 10.1136/bmjment-2024-301424

Hawks, Z. W., & Constantino, J. N. (2020). Neuropsychiatric “Comorbidity” as Causal Influence in Autism. J Am Acad Child Adolesc Psychiatry, 59(2), 229–235. 10.1016/j.jaac.2019.07.008

Hirota, T., & King, B. H. (2023). Autism Spectrum Disorder: A Review. JAMA, 329(2), 157–168. 10.1001/jama.2022.23661

Hus, V., & Lord, C. (2014). The autism diagnostic observation schedule, module 4: revised algorithm and standardized severity scores. Journal of autism and developmental disorders, 44, 1996–2012.

Jiujias, M., Kelley, E., & Hall, L. (2017). Restricted, Repetitive Behaviors in Autism Spectrum Disorder and Obsessive-Compulsive Disorder: A Comparative Review. Child Psychiatry Hum Dev, 48(6), 944–959. 10.1007/s10578-017-0717-0

Kaufman, A. S., Kaufman, N. L. (2004). Kaufman Brief Intelligence Test, Second Edition. American Guidance Services.

Kavanaugh, B. C., Gabert, T., Jones, R. N., Sheinkopf, S. J., & Morrow, E. M. (2022). Parental age and autism severity in the Rhode Island Consortium for Autism Research and Treatment (RI-CART) study. Autism Res, 15(1), 86–92. 10.1002/aur.2648

Kavanaugh, B. C., Schremp, C. A., Jones, R. N., Best, C. R., Sheinkopf, S. J., & Morrow, E. M. (2021). Moderators of Age of Diagnosis in > 20,000 Females with Autism in Two Large US Studies. J Autism Dev Disord. 10.1007/s10803-021-05026-4

Knott, R., Mellahn, O. J., Tiego, J., Kallady, K., Brown, L. E., Coghill, D., Williams, K., Bellgrove, M. A., & Johnson, B. P. (2024). Age at diagnosis and diagnostic delay across attention-deficit hyperactivity and autism spectrums. Aust N Z J Psychiatry, 58(2), 142–151. 10.1177/00048674231206997

Leng, L. L., Zhu, Y. W., & Zhou, L. G. (2024). Explaining differences in autism detection timing: Age of diagnosis and associated individual and socio-familial factors in Chinese children. Autism, 28(4), 896–907. 10.1177/13623613231187184

Lord, C., Elsabbagh, M., Baird, G., & Veenstra-Vanderweele, J. (2018). Autism spectrum disorder. The Lancet, 392(10146), 508–520. 10.1016/s0140-6736(18)31129-2

Lord, C., Rutter, M., DiLavore, P. C., Risi, S., Gotham, K., & Bishop, S. (2012). Autism Diagnostic Observation Schedule, Second Edition. Western Psychological Services.

Loubersac, J., Michelon, C., Ferrando, L., Picot, M. C., & Baghdadli, A. (2023). Predictors of an earlier diagnosis of Autism Spectrum Disorder in children and adolescents: a systematic review (1987-2017). Eur Child Adolesc Psychiatry, 32(3), 375–393. 10.1007/s00787-021-01792-9

Mandy, W., Midouhas, E., Hosozawa, M., Cable, N., Sacker, A., & Flouri, E. (2022). Mental health and social difficulties of late-diagnosed autistic children, across childhood and adolescence. J Child Psychol Psychiatry, 63(11), 1405–1414. 10.1111/jcpp.13587

Mannion, A., & Leader, G. (2013). Comorbidity in autism spectrum disorder: A literature review. Research in Autism Spectrum Disorders, 7(12), 1595–1616. 10.1016/j.rasd.2013.09.006

McCormick, C. E. B., Kavanaugh, B. C., Sipsock, D., Righi, G., Oberman, L. M., Moreno De Luca, D., Gamsiz Uzun, E. D., Best, C. R., Jerskey, B. A., Quinn, J. G., Jewel, S. B., Wu, P. C., McLean, R. L., Levine, T. P., Tokadjian, H., Perkins, K. A., Clarke, E. B., Dunn, B., Gerber, A. H., . . . Morrow, E. M. (2020). Autism Heterogeneity in a Densely Sampled U.S. Population: Results From the First 1,000 Participants in the RI-CART Study. Autism Res, 13(3), 474–488. 10.1002/aur.2261

Memisevic, H., Pasalic, A., & Saletovic, A. (2023). Autism severity level affects working memory and planning but not inhibition, shifting and emotional control. Autism Res, 16(7), 1335–1343. 10.1002/aur.2952

Nordahl, C. W., Andrews, D. S., Dwyer, P., Waizbard-Bartov, E., Restrepo, B., Lee, J. K., Heath, B., Saron, C., Rivera, S. M., Solomon, M., Ashwood, P., & Amaral, D. G. (2021). The Autism Phenome Project: Toward Identifying Clinically Meaningful Subgroups of Autism. Front Neurosci, 15, 786220. 10.3389/fnins.2021.786220

Ochoa-Lubinoff, C., Makol, B. A., & Dillon, E. F. (2023). Autism in Women. Neurol Clin, 41(2), 381–397. 10.1016/j.ncl.2022.10.006

Rattaz, C., Loubersac, J., Michelon, C., Geoffray, M. M., Picot, M. C., Munir, K., & Baghdadli, A. (2022). Factors associated with age of diagnosis in children with autism spectrum disorders: Report from a French cohort. Autism, 26(8), 2108–2116. 10.1177/13623613221077724

Rutter, M., Bailey, A., & Lord, C. (2003). Social communication questionnaire. Western Psychological Services.

Sainsbury, W. J., Carrasco, K., Whitehouse, A. J. O., & Waddington, H. (2023). Parent-reported Early Atypical Development and Age of Diagnosis for Children with Co-occurring Autism and ADHD. J Autism Dev Disord, 53(6), 2173–2184. 10.1007/s10803-022-05488-0

Simonoff, E., Pickles, A., Charman, T., Chandler, S., Loucas, T., & Baird, G. (2008). Psychiatric disorders in children with autism spectrum disorders: prevalence, comorbidity, and associated factors in a population-derived sample. Journal of the American Academy of Child & Adolescent Psychiatry, 47(8), 921–929.

Soke, G. N., Maenner, M. J., Christensen, D., Kurzius-Spencer, M., & Schieve, L. A. (2018). Prevalence of Co-occurring Medical and Behavioral Conditions/Symptoms Among 4- and 8-Year-Old Children with Autism Spectrum Disorder in Selected Areas of the United States in 2010. J Autism Dev Disord, 48(8), 2663–2676. 10.1007/s10803-018-3521-1

SPARKConsortium. (2018). SPARK: A US Cohort of 50,000 Families to Accelerate Autism Research. Neuron, 97(3), 488–493. 10.1016/j.neuron.2018.01.015

Sparrow, S., Balla, D., Cicchetti, D. (2005). Vineland Adaptive Behavior Scales. American Guidance Service.

Zuckerman, K., Lindly, O. J., & Chavez, A. E. (2017). Timeliness of Autism Spectrum Disorder Diagnosis and Use of Services Among U.S. Elementary School-Aged Children. Psychiatr Serv, 68(1), 33–40. 10.1176/appi.ps.201500549

Zwaigenbaum, L., Bauman, M., Choueiri, R., Kasari, C., Carter, A., Granpeesheh, D., Mailloux, Z., Roley, S. S., Wagner, S., Fein, D., Pierce, K., Buie, T., Davis, P. A., Newschaffer C., Robins, D., Wetherby, A., Stone, W. L., Yirmiya, N., Estes, A., Hansen, R., McPartland, J. C., Natowicz, M. R. (2015). Early intervention for children with autism spectrum disorder under 3 years of age: Recommendations for practice and research. Pediatrics.

